# The impacts of COVID-19 vaccine timing, number of doses, and risk prioritization on mortality in the US

**DOI:** 10.1101/2021.01.18.21250071

**Authors:** Xutong Wang, Zhanwei Du, Kaitlyn E. Johnson, Remy F. Pasco, Spencer J. Fox, Michael Lachmann, Jason S. McLellan, Lauren Ancel Meyers

**Affiliations:** Department of Integrative Biology, The University of Texas at Austin, Austin, Texas, United States of America; WHO Collaborating Centre for Infectious Disease Epidemiology and Control, School of Public Health, LKS Faculty of Medicine, The University of Hong Kong, Hong Kong Special Administrative Region, China; Laboratory of Data Discovery for Health, Hong Kong Science and Technology Park, Hong Kong Special Administrative Region, China; Santa Fe Institute, Santa Fe, New Mexico, United States of America; Department of Molecular Biosciences, The University of Texas at Austin, Austin, Texas, United States of America

## Abstract

As COVID-19 vaccination begins worldwide, policymakers face critical trade-offs. Using a mathematical model of COVID-19 transmission, we find that timing of the rollout is expected to have a substantially greater impact on mortality than risk-based prioritization and uptake and that prioritizing first doses over second doses may be life saving.

## Text

In December 2020, the US government issued emergency use authorization for two-dose SARS-CoV-2 vaccines manufactured by Moderna and Pfizer-BioNTech, both estimated to be over 94% efficacious in preventing symptomatic COVID-19 infections (1–3). The Advisory Committee on Immunization Practices immediately recommended the prioritization of front-line workers and high-risk subgroups (4). As of February 14, roughly 52 million doses have been administered (5). We used a mathematical model of COVID-19 transmission to evaluate the impact of vaccine timing, risk prioritization, numbers of doses administered and rates of uptake on population-level mortality (Figure).

Focusing on Austin, Texas, we project COVID-19 deaths over eight months for both an infection-blocking vaccine that prevents infection upon exposure and a symptom-blocking vaccine that prevents symptoms upon infection, assuming all vaccinated individuals enjoy a 95% reduction in susceptibility (i.e., a *leaky* vaccine). Vaccination begins on either January 15 or February 15, with 10M vaccines administered weekly and allocated to cities pro rata (Figure A1.1). We compare three strategies: no priority groups; one of three priority groups vaccinated prior to the general public––adults over 65, adults with high-risk comorbidities, or both of these groups; ten phases that vaccinate age-risk groups in order of risk of severe COVID-19 outcomes. Stochastic simulations assume that 7.6% of individuals have been immunized by infection prior to January 15th.

If a perfectly risk-prioritized (ten-phase) rollout of an infection-blocking vaccine begins January 15, we estimate that 52% (95% CI: 47% - 56%) or 56% (95% CI: 51% - 60%) of the deaths would be averted relative to the baseline of no vaccines, assuming 50% or 90% uptake, respectively (Figure). If delayed by one month, 34% (95% CI: 28%-40%) or 38% (95% CI: 32%-43%) of deaths would be averted, respectively. Under low (50%) uptake, prioritization has minimal benefit. Under high uptake (90%), the ten-stage strategy is optimal, followed by prioritizing everyone over 65 and high-risk young adults.

If the vaccine is only symptom-blocking, then the expected differences are magnified. With a January 15 start and 50% uptake, the ten-phase strategy is expected to avert 40% (95% CI: 35%-45%) of deaths whereas the unprioritized rollout would avert 32% (95% CI: 25%-37%). If only a single dose with 80% efficacy (1,2) is administered under the ten-phase strategy, we would expect a 50% (95% CI: 45%-54%) reduction in mortality for a symptom-blocking vaccine and 66% (95% CI: 63%-70%) reduction for an infection-blocking vaccine (Table A1.1).

These projections validate the prioritization of high-risk groups. Under a pessimistic scenario in which a symptom-blocking vaccine is rolled out starting February 2021 with only 50% uptake, prioritizing high risk adults and those over 65 is expected to avert ∼17,000 (95% CI: 0-36,000) more deaths in the US than a non-prioritized campaign. However, given the alarming state of the pandemic in early 2021, with thousands dying daily across the US and Europe, vaccine delays are expected to cost more lives than either imperfect prioritization or vaccine hesitancy. Unlike seasonal influenza vaccination campaigns, which begin prior to the widespread transmission of the virus, the global health community is racing to get COVID-19 vaccines to people before the virus reaches them. With the emergence of potentially more contagious variants in the UK and South Africa, this may become even more challenging.

The UK and Belgium have prioritized first doses over second doses (6), in an effort to provide partial immunity to more people. The US has publicly resisted this approach, citing the lack of clinical trial data validating the approach (7). We find that providing a single (80% efficacious) dose would be expected to save more lives than the corresponding two-dose strategy, because partially immunizing a large number of people confers a greater degree of population-level protection than more fully immunizing half as many. Although a one-dose campaign may accelerate herd immunity and require far fewer resources than a two-dose campaign, we strongly caution that additional data and single-dose trials are needed to establish efficacy and note that low-efficacy vaccines may increase the risk of vaccine-resistant variants and that there may be political, commercial and societal barriers to shifting priorities mid-campaign (9).

We assume that vaccines provide lasting immunity and block either infection or symptoms, whereas the reality may be a hybrid of the two (Figure A1.2 and Table A1.2), and do not consider riskier behavior stemming from weariness or overconfidence in the vaccination campaign. Our estimates also reflect conditions in the US in early 2021, with cases surging toward a pandemic peak within months in the absence of effective mitigation. The estimated public health benefits of vaccines decrease under higher COVID-19 transmission rates that might occur with the relaxation of mitigation measures, lower levels of immunity prior to the rollout, or the emergence of more transmissible SARS-CoV-2 variants including B.1.1.7 (Appendix).

Risk prioritization is a valid approach for maximizing the impact of vaccines, but not at the expense of vaccination speed. Our projections suggest two immediate strategies to amplify the impact of COVID-19 vaccines in the US––hybrid distributions that combine active outreach to priority groups with passive distribution to the general public and foregoing plans to hold second doses in reserve.

**Figure.**
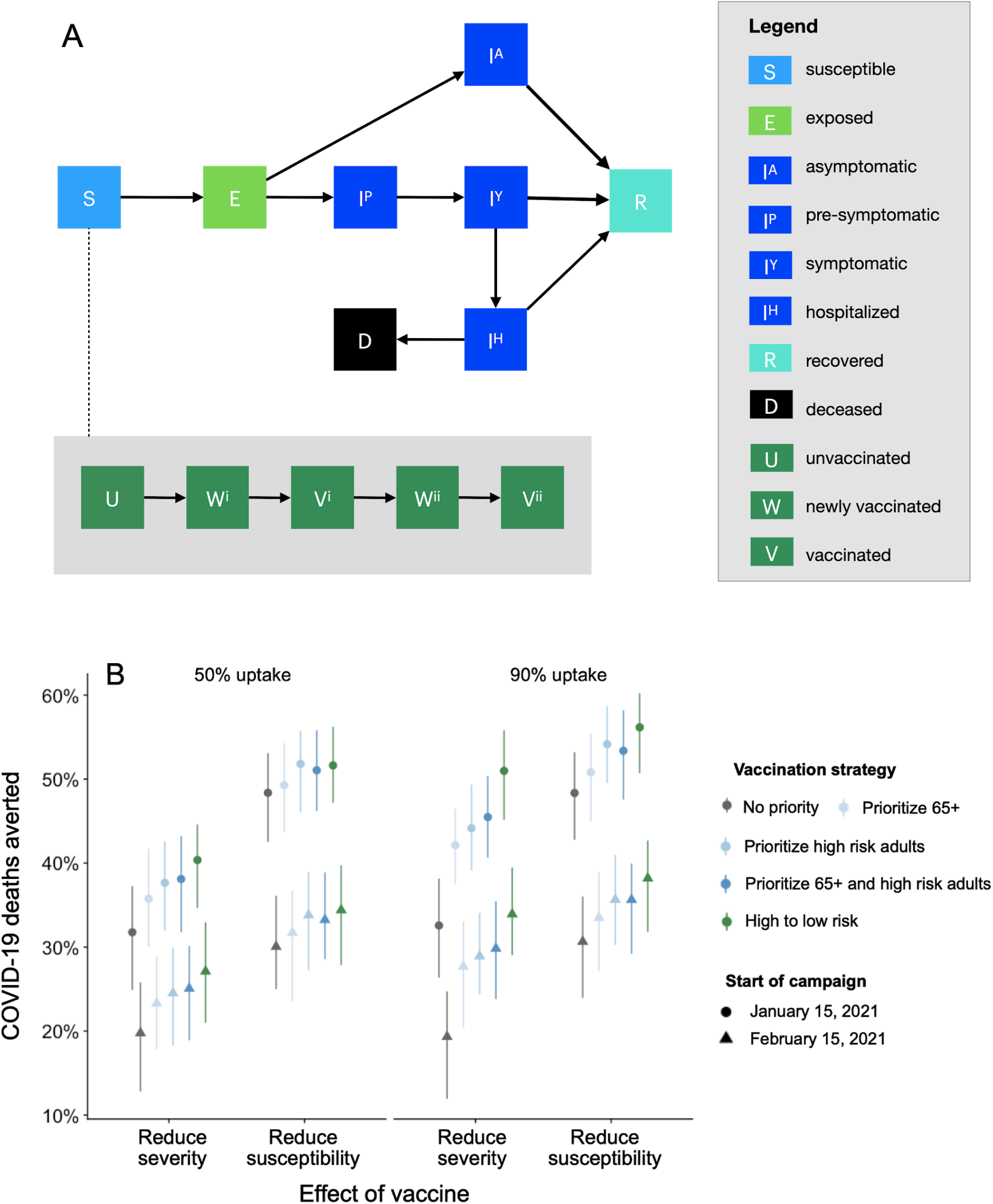
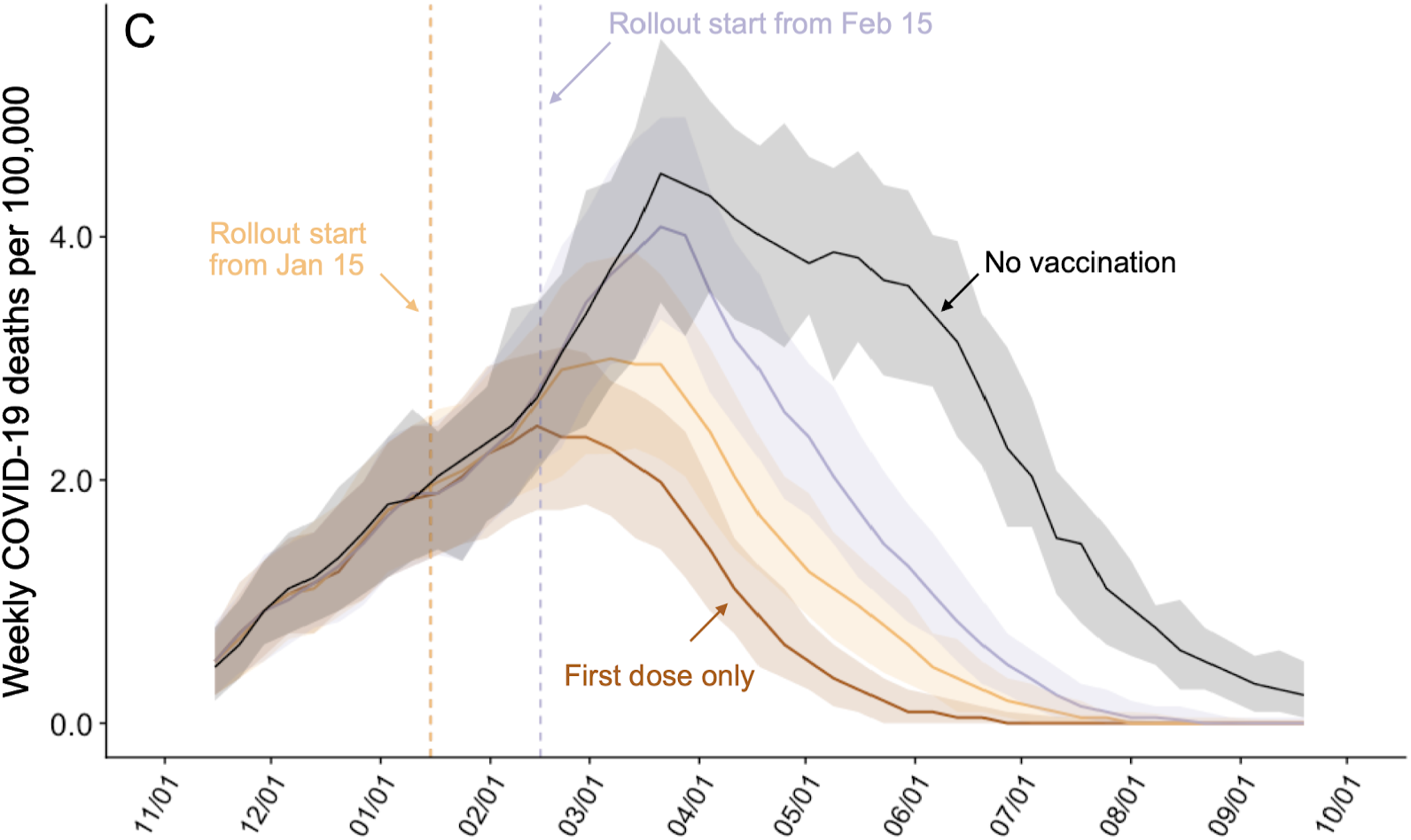
Model structure and projected COVID-19 mortality in the Austin-Round Rock MSA from November 8, 2020 to September 17, 2021 under various vaccine rollout scenarios. A) Compartmental model of COVID-19 transmission in a US city. Each age-risk subgroup is modeled with a separate set of compartments. Upon infection, susceptible individuals (*S*) progress to exposed (*E*) and then to either pre-symptomatic infectious (*I*^*P*^) or asymptomatic infectious (*I*^*A*^). All asymptomatic cases eventually progress to a recovered class where they remain protected from future infection (*R*); pre-symptomatic cases progress to symptomatic (*I*^*Y*^) then are either hospitalized (*I*^*H*^) or recover. Mortality (*D*) varies by age group and risk group and is assumed to be preceded by hospitalization. Within each compartment, individuals are divided by vaccination status: unvaccinated (*U*), newly vaccinated with the first dose (*W*^*i*^), vaccinated with the first dose (*V*^*i*^), newly vaccinated with the second dose (*W*^*ii*^), and fully vaccinated with the second dose (*V*^*ii*^). We model stochastic transitions between compartments and sample vaccine efficacy parameters from distributions to capture uncertainty, while keeping all other parameters fixed (Appendix). B) COVID-19 deaths averted after January 15, 2021 under combinations of: vaccine uptake, either 50% (left) or 90% (right); type of protection, either infection or symptom blocking (x-axis); rollout dates, either January 15 (circles) or February 15 (triangles); and risk prioritization, either no priority (gray), prioritize all adults over 65y (light blue), adults with high-risk comorbidities (medium blue), or the combination of the two (dark blue), or a ten-phase risk-ordered strategy (green) that sequentially vaccinates >65y high risk, 50-64y high risk, >65y low risk, 18-49y high risk, 50-64y low risk, 18-49y low risk, 0-4y high risk, 5-17y high risk, 0-4y low risk, 5-17y low risk. Points and whiskers indicate the median and 95% CI across 200 paired stochastic simulations. C) Weekly incident COVID-19 deaths per 100,000 assuming intermediate (70%) uptake (10) without vaccine (black) or under a ten-phase risk-based rollout of a 95% efficacious infection-blocking, starting either January 15 (orange) or February 15 (purple). The brown line assumes that only first doses are administered starting January 15. Solid lines and shading indicate the median and 95% CI across 200 stochastic simulations, respectively.

## Data Availability

NA

## Acknowledgements

We acknowledge the critical discussions and parameter guidance from Matthew Biggerstaff at the US CDC. This research was supported by NIH Grant R01 AI151176, CDC Grant U01 IP00136, and a donation from Love, Tito’s (the philanthropic arm of Tito’s Homemade Vodka, Austin, TX, USA) to the University of Texas to support the modeling of COVID-19 mitigation strategies.

## Author Bio

Xutong Wang is a Ph.D. candidate at the University of Texas at Austin under the supervision of Dr. Lauren Ancel Meyers. Her research interest is on mathematical and statistical modeling of infectious disease dynamics.

