## Appendix for "The impacts of COVID-19 vaccine timing, number of doses, and risk prioritization on mortality in the US"

**Affiliations:**

### Supplementary Material

#### Section 1. Supplementary Figures and Tables

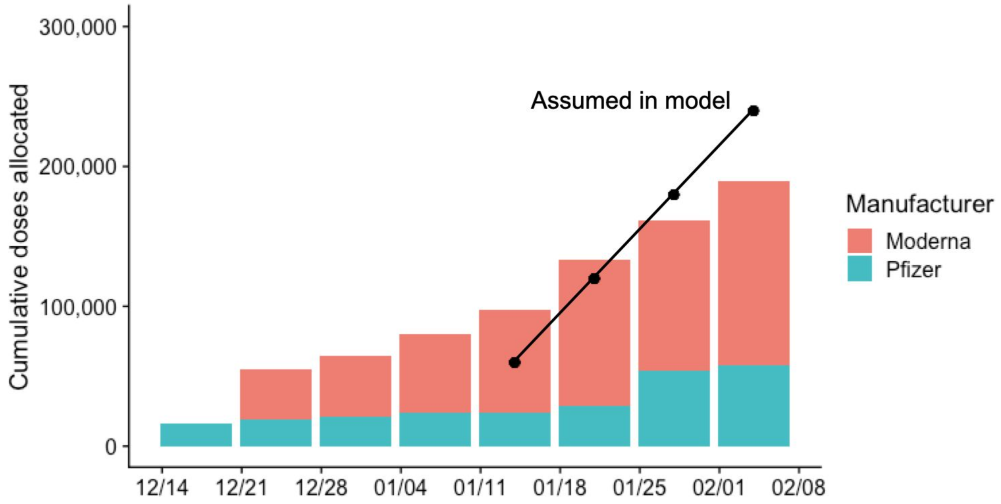


**Figure A1.1. COVID-19 vaccine doses allocated to the Austin-Round Rock MSA in comparison to the model assumption** (1)**.** Red represents doses manufactured by Moderna and green represents vaccines manufactured by Pfizer-BioNTech. Black points indicate the rollout assumed by the model.

**Table A1.1. Percent of COVID-19 deaths after January 15, 2021 averted.** Values are medians and 95% confidence intervals based on 200 pairs of stochastic simulations. Deaths averted are computed by comparing a simulation of the specified vaccine strategy to a simulation without vaccination.

| Vaccine rollout date | | January 15 | | | February 15 | | |
|---|---|---|---|---|---|---|---|
| Vaccine uptake | | 0.5 | 0.7 | 0.9 | 0.5 | 0.7 | 0.9 |
| Infection blocking 2 dose | All | 48 (43-53) | 48 (42-54) | 48 (43-53) | 30 (25-36) | 30 (25-35) | 31 (24 -36) |
| | >65 | 49 (44-54) | 50 (45-55) | 51 (45-55) | 32 (24-37) | 33 (27-38) | 33 (27-39) |
| | high risk adults | 52 (46-56) | 54 (48-58) | 54 (50-59) | 34 (27-39) | 35 (30-40) | 36 (30-41) |
| | >65 + high risk | 51 (46-56) | 52 (48-58) | 53 (48-58) | 33 (29-39) | 34 (27-40) | 36 (29-40) |
| | 10 phase | 52 (47-56) | 54 (49-58) | 56 (51-60) | 34 (28-40) | 36 (30-42) | 38 (32-43) |
| Symptom blocking 2 dose | No priorities | 32 (25-37) | 32 (27-38) | 33 (26-38) | 20 (13-26) | 19 (13-25) | 19 (12-25) |
| | >65 | 36 (30-42) | 40 (34-44) | 42 (38-46) | 23 (18-29) | 26 (19-32) | 28 (20-33) |
| | high risk adults | 38 (32-43) | 41 (35-47) | 44 (39-49) | 25 (18-30) | 27 (21-33) | 29 (24-34) |
| | >65 + high risk | 38 (32-43) | 43 (38-49) | 45 (41-50) | 25 (19-30) | 28 (22-33) | 30 (24-35) |
| | 10 phase | 40 (35-45) | 46 (40-51) | 51 (45-56) | 27 (21-33) | 32 (25-37) | 34 (29-39) |
| Infection blocking 1 dose | No priorities | 65 (61-68) | 65 (62-69) | 65 (63-69) | 44 (39-49) | 44 (40-49) | 44 (40-50) |
| | >65 | 66 (62-69) | 66 (63-70) | 68 (63-71) | 45 (41-50) | 47 (42-52) | 47 (43-52) |
| | high risk adults | 66 (62-69) | 68 (64-72) | 70 (66-73) | 46 (41-52) | 48 (43-53) | 49 (43-54) |
| | >65 + high risk | 66 (63-70) | 68 (64-71) | 69 (65-73) | 46 (41-51) | 48 (43-53) | 49 (44-54) |
| | 10 phase | 66 (63-70) | 69 (66-73) | 71 (67-74) | 47 (41-51) | 49 (44-54) | 51 (46-55) |
| Symptom blocking 1 dose | No priorities | 46 (39-50) | 50 (45-55) | 51 (47-56) | 31 (25-36) | 33 (27-39) | 34 (28-38) |
| | >65 | 48 (42-52) | 54 (49-58) | 57 (52-61) | 33 (28-38) | 37 (32-43) | 39 (33-45) |
| | high risk adults | 49 (43-53) | 55 (49-60) | 58 (54-63) | 34 (29-39) | 38 (33-43) | 41 (35-45) |
| | >65 + high risk | 49 (43-53) | 55 (51-60) | 60 (55-64) | 34 (28-39) | 39 (33-43) | 42 (36-46) |
| | 10 phase | 50 (45-54) | 57 (53-62) | 63 (58-67) | 35 (29-40) | 41 (36-46) | 45 (39-50) |

#### Hybrid scenarios: Infection-blocking and symptom-blocking vaccine

Our model assumes the vaccine is either infection blocking or symptom blocking, while the reality may be somewhere in between. Therefore, we projected the COVID-19 mortality under a hybrid scenario where two doses of the vaccine are 67% efficacious against all infections and 82% efficacious against developing symptoms, with additional parameters given in Table A2.4 (Figure A1 and Table A1.2).

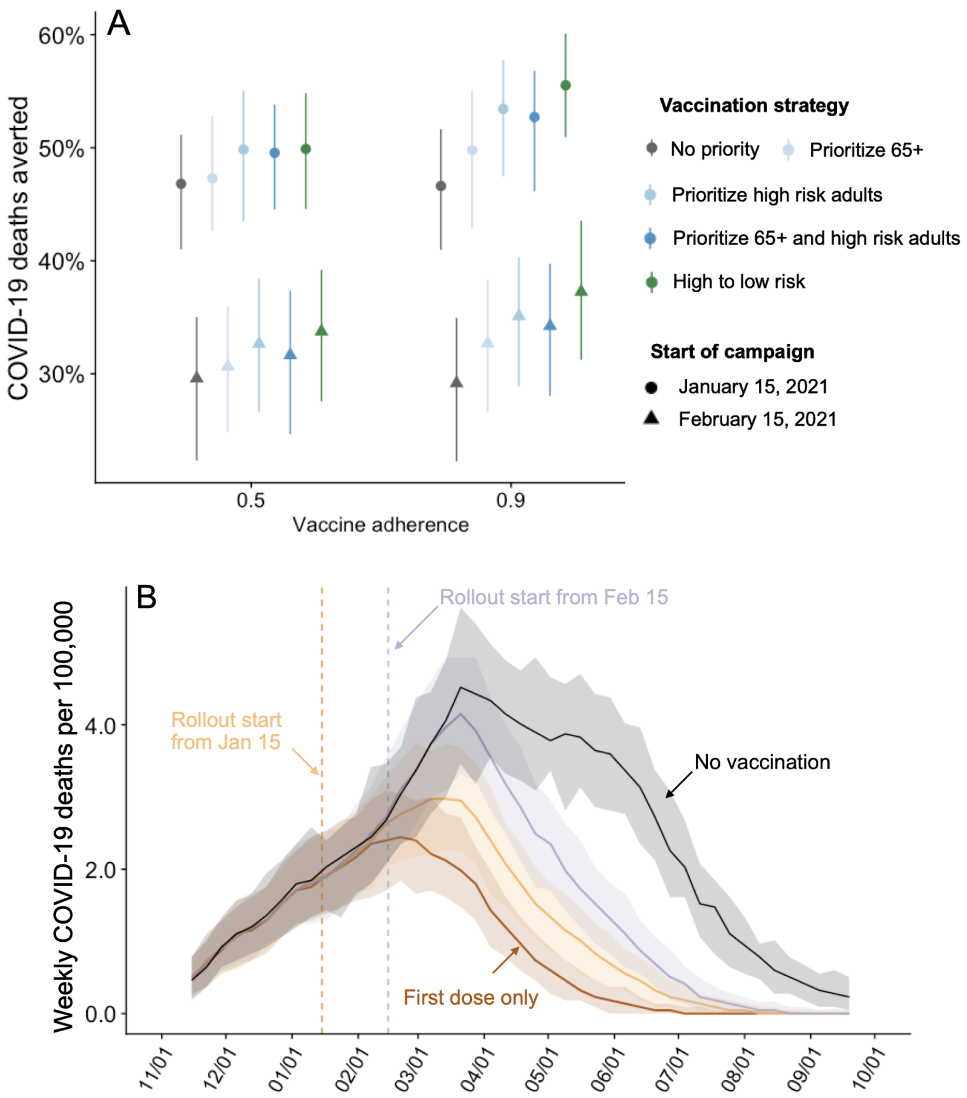


**Figure A1.2. Projected COVID-19 mortality in the Austin-Round Rock MSA from November 8, 2020 to September 17, 2021 under various vaccine rollout scenarios.** A) COVID-19 deaths averted after January 15, 2021 for a hybrid type of vaccination protection under combinations of: vaccine uptake, either 50% or 90% (x-axis); rollout dates, either January 15 (circles) or February 15 (triangles); and risk prioritization, either no priority (gray), prioritize all adults over 65y (light blue), adults with high-risk comorbidities (medium blue), or the combination of the two (dark blue), or a ten-phase risk-ordered strategy (green) that sequentially vaccinates >65y high risk, 50-64y high risk, >65y low risk, 18-49y high risk, 50-64y low risk, 18-49y low risk, 0-4y high risk, 5-17y high risk, 0-4y low risk, 5-17y low risk. Points and whiskers indicate the median and 95% CI across 200 paired stochastic simulations. B) Weekly incident COVID-19 deaths per 100,000 assuming intermediate (70%) uptake (2) without vaccine (black) or under a ten-phase risk-based rollout of a 95% efficacious infection-blocking, starting either January 15 (orange) or February 15 (purple). The brown line assumes that only first doses are administered starting January 15. Solid lines and shading indicate the median and 95% CI across 200 stochastic simulations, respectively.

**Table A1.2. Percent of COVID-19 deaths after January 15, 2021 averted for hybrid scenarios.** Values are medians and 95% confidence intervals based on 200 pairs of stochastic simulations. Deaths averted are computed by comparing a simulation of the specified vaccine strategy to a simulation without vaccination.

| **Vaccine rollout date** | | **January 15** | | | **February 15** | | |
|---|---|---|---|---|---|---|---|
| **Vaccine uptake** | | **0.5** | **0.7** | **0.9** | **0.5** | **0.7** | **0.9** |
| **Hybrid 2 dose** | **No priorities** | 47 (41-51) | 46 (41-51) | 47 (41-52) | 30 (22-35) | 29 (23-35) | 29 (22-35) |
| | **>65** | 47 (43-53) | 48 (43-54) | 50 (43-55) | 31 (25-36) | 32 (25-37) | 33 (27-38) |
| | **high risk adults** | 50(43-55) | 52(46-57) | 53(47-58) | 33(27-38) | 34(27-39) | 35(29-40) |
| | **>65 + high risk** | 50 (45-54) | 51 (46-56) | 53 (46-57) | 32 (25-37) | 34 (27-39) | 34 (28-40) |
| | **10 phase** | 50 (45-55) | 53 (48-58) | 56 (51-60) | 34 (28-39) | 36 (30-41) | 37 (31-44) |
| **Hybrid 1 dose** | **No priorities** | 63 (59-67) | 64 (60-68) | 64 (59-67) | 43 (37-47) | 43 (37-48) | 44 (38-48) |
| | **>65** | 64 (59-68) | 65 (61-69) | 66 (62-70) | 44 (39-49) | 45 (40-50) | 46 (41-52) |
| | **high risk adults** | 65 (61-68) | 67 (62-70) | 68 (65-72) | 45 (39-49) | 47 (41-52) | 48 (43-52) |
| | **>65 + high risk** | 64 (60-68) | 67 (63-70) | 68 (64-71) | 45 (40-50) | 47 (42-51) | 48 (44-52) |
| | **10 phase** | 65 (61-69) | 68 (64-72) | 70 (66-73) | 46 (40-50) | 48 (44-53) | 50 (46-54) |

### Section 2. Stochastic Compartmental Model of COVID-19 Transmission in the Austin-Round Rock Metropolitan Area

The main Figure includes a diagram of the model structure. For each vaccine, age and risk group, we build a separate set of compartments to model the transitions between the states: susceptible ($S$), exposed ($E$), pre-symptomatic infectious ($I^P$), symptomatic infectious ($I^Y$), asymptomatic infectious ($I^A$), symptomatic infectious that are hospitalized ($I^H$), recovered ($R$), and deceased ($D$). The symbols $S$, $E$, $I^P$, $I^Y$, $I^A$, $I^H$, $R$, and $D$ denote the number of people in that state in the given vaccine/age/risk group and the total size of the vaccine/age/risk group is $N = S + E + I^P + I^Y + I^A + I^H + R + D$.

The model for individuals in vaccine group $v$, age group $a$, and risk group $r$ is given by:

$$\frac{dS_{v,a,r}}{dt} = -\sum_{u \in V} \sum_{i \in A} \sum_{j \in K} S_{v,a,r} \kappa_{v,a,r} \beta_0 \phi_{a,i} \left( \omega_u^P I_{u,i,j}^P + \omega_u^A I_{u,i,j}^A + \omega_u^Y I_{u,i,j}^Y \right) / N_i$$

$$\frac{dE_{v,a,r}}{dt} = \sum_{u \in V} \sum_{i \in A} \sum_{j \in K} S_{v,a,r} \kappa_{v,a,r} \beta_0 \phi_{a,i} \left( \omega_u^P I_{u,i,j}^P + \omega_u^A I_{u,i,j}^A + \omega_u^Y I_{u,i,j}^Y \right) / N_i - \sigma E_{v,a,r}$$

$$\frac{dI_{v,a,r}^A}{dt} = (1 - \lambda_{v,a,r} \tau) \sigma E_{v,a,r} - \gamma^A I_{v,a,r}^A$$

$$\frac{dI^P_{v,a,r}}{dt} = \lambda_{v,a,r}\tau\sigma E_{v,a,r} - \rho I^P_{v,a,r}$$

$$\frac{dI^Y_{v,a,r}}{dt} = \rho I^P_{v,a,r} - (1-\pi)\gamma^Y I^Y_{v,a,r} - \pi\eta I^Y_{v,a,r}$$

$$\frac{dI^H_{v,a,r}}{dt} = \pi\eta I^Y_{v,a,r} - (1-\nu)\gamma^H I^H_{v,a,r} - \nu\mu I^H_{v,a,r}$$

$$\frac{dR_{v,a,r}}{dt} = \gamma^A I^A_{v,a,r} + (1-\pi)\gamma^Y I^Y_{v,a,r} + (1-\nu)\gamma^H I^H_{v,a,r}$$

$$\frac{dD_{v,a,r}}{dt} = \nu\mu I^H_{v,a,r}$$

where $V$, $A$ and $K$ are all possible vaccine, age and risk groups, $\omega^A$, $\omega^Y$, $\omega^P$ are relative infectiousness of the $I^A$, $I^Y$, $I^P$ compartments, respectively, $\beta_0$ is baseline transmission rate, $\phi_{a,i}$ is the mixing rate between age group $a$, $i \in A$, $\gamma^A, \gamma^Y$, $\gamma^H$ are the recovery rates for the $I^A$, $I^Y$, $I^H$ compartments, respectively, $\sigma$ is the exposed rate, $\tau$ is the symptomatic ratio, $\rho$ is the rate from pre-symptomatic to symptomatic, $\pi$ is the proportion of symptomatic individuals requiring hospitalization, $\eta$ is rate at which hospitalized cases enter the hospital following symptom onset, $\nu$ is mortality rate for hospitalized cases, and $\mu$ is rate at which terminal patients die.

The transition between each vaccine group $v$, is given by

$$\frac{dX_{U,a,r}}{dt} =- \alpha^i_{a,r} X_{U,a,r}$$

$$\frac{dX_{W^i,a,r}}{dt} = \alpha^i_{a,r} X_{U,a,r} - \delta^i X_{W^i,a,r}$$

$$\frac{dX_{V^i,a,r}}{dt} = \delta^i X_{W^i,a,r} - \alpha^{ii}_{a,r}\delta X_{V^i,a,r}$$

$$\frac{dX_{W^{ii},a,r}}{dt} = \alpha^{ii}_{a,r}\delta X_{V^i,a,r} - \delta^{ii} X_{W^{ii},a,r}$$

$$\frac{dX_{V^{ii},a,r}}{dt} = \delta^{ii} X_{W^{ii},a,r}$$

where $X \in \{S, E, I^A, I^P, I^Y, I^H, R, D\}$, and $v \in \{U, W^i, V^i, W^{ii}, V^{ii}\}$, $\alpha^i$ is vaccination rate, $\delta^i$ is the rate of gaining partial immunity after the first dose, $\alpha^{ii}$ is the proportion of individuals whole return for a second dose after receiving the first dose (i.e., the second dose return rate), $\delta$

is the rate of obtaining the second dose following the first dose (for those who return), and $\delta^{ii}$ is the rate of gaining immunity after the second dose.

We model stochastic transitions between compartments using the $\tau$-leap method (3,4) with key parameters given in Table A2. Assuming that the events at each time-step are independent and do not impact the underlying transition rates, the numbers of each type of event should follow Poisson distributions with means equal to the rate parameters. We thus simulate the model according to the following equations:

$$S_{v,a,r}(t+1) - S_{v,a,r}(t) = -P_1$$

$$E_{v,a,r}(t+1) - E_{v,a,r}(t) = P_1 - P_2$$

$$I^A_{v,a,r}(t+1) - I^A_{v,a,r}(t) = (1 - \lambda_{v,a,r}\tau) P_2 - P_3$$

$$I^P_{v,a,r}(t+1) - I^P_{v,a,r}(t) = \lambda_{v,a,r}\tau P_2 - P_4$$

$$I^Y_{v,a,r}(t+1) - I^Y_{v,a,r}(t) = P_4 - P_5 - P_6$$

$$I^H_{v,a,r}(t+1) - I^H_{v,a,r}(t) = P_6 - P_7 - P_8$$

$$D_{v,a,r}(t+1) - D_{v,a,r}(t) = P_7$$

$$R_{v,a,r}(t+1) - R_{v,a,r}(t) = P_3 + P_5 + P_8$$

$$X_{U,a,r}(t+1) - X_{U,a,r}(t) = -\alpha^i_{a,r}(t) X_{U,a,r}(t)$$

$$X_{W^i,a,r}(t+1) - X_{W^i,a,r}(t) = \alpha^i_{a,r}(t) X_{U,a,r}(t) - P_9$$

$$X_{V^i,a,r}(t+1) - X_{V^i,a,r}(t) = P_9 - P_{10}$$

$$X_{W^{ii},a,r}(t+1) - X_{W^{ii},a,r}(t) = P_{10} - P_{11}$$

$$X_{V^{ii},a,r}(t+1) - X_{V^{ii},a,r}(t) = P_{11}$$

with

$$P_1 \sim Pois(S_{v,a,r}(t) F_{v,a,r}(t))$$

$$P_2 \sim Pois(\sigma E_{v,a,r}(t))$$

$$P_3 \sim Pois\left(\gamma^A I^A_{v,a,r}(t)\right)$$

$$P_4 \sim Pois\left(\rho I^P_{v,a,r}(t)\right)$$

$$P_5 \sim Pois\left((1-\pi)\gamma^Y I^Y_{v,a,r}(t)\right)$$

$$P_6 \sim Pois\left(\pi\eta I^Y_{v,a,r}(t)\right)$$

$$P_7 \sim Pois\left(\nu\mu I^H_{v,a,r}(t)\right)$$

$$P_8 \sim Pois\left((1-\nu)\gamma^H I^H_{v,a,r}(t)\right)$$

$$P_9 \sim Pois\left(\delta^i X_{W^i,a,r}(t)\right)$$

$$P_{10} \sim Pois\left(\alpha^{ii}\delta X_{V^i,a,r}(t)\right)$$

$$P_{11} \sim Pois\left(\delta^{ii} X_{W^{ii},a,r}(t)\right)$$

and where $F_{v,a,r}$ denotes the force of infection for individuals in vaccine group *v*, age group *a*, and risk group *r* and is given by:

$$F_{v,a,r}(t) = \sum_{u\in V}\sum_{i\in A}\sum_{j\in K} \kappa_{v,a,r}\beta_0\phi_{a,i}\left(\omega_u^P I^P_{u,i,j} + \omega_u^A I^A_{u,i,j} + \omega_u^Y I^Y_{u,i,j}\right)/N_i\,.$$

#### Model Parameters

**Table A2.1. Initial conditions, school calendar and contact rates**

| **Variable** | **Settings** |
|---|---|
| Initial day of simulation | 11/8/2020 |
| Initial number infected | Based on estimates for Austin, Texas, given Table A2.9 (5) |
| Age-specific and day-specific contact rates[a] | Home, work, other and school matrices provided in Tables A2.5-A2.8<br>Typical weekday = home + work + other + school<br>Weekends and holiday weekdays= home + other<br>Weekdays during non-holiday school breaks = home + work + other |
| School calendar | Austin Independent School District calendar (2019-2020, 2020-2021) (6) |

[a] We assume the age-specific contact rates given in ref. (7), which takes the contact numbers estimated through diary-based POLYMOD study in Europe (8) and extrapolates to the United States. The values in Tables A2.5-A2.8 are the assumed daily contacts between each pair of age groups at home, school, work, and all other places, respectively. These contact matrices are used to adjust the transmission rate between age groups. The accuracy of the contact matrices is limited by (i) possible biases with the original diary-based study (8), (ii) assumptions made when projecting the original study to the US (7), and (iii) impacts of COVID-19 policies and perceptions on daily contact patterns.

**Table A2.2. Epidemiological parameters.** Values given as five-element vectors are age-stratified with values corresponding to 0-4, 5-17, 18-49, 50-64, 65+ year age groups, respectively.

| **Parameters** | **Best guess values** | **Source** |
|---|---|---|
| $R_e$: effective reproduction number | 1.2 | (5) |
| $\beta$ : baseline transmission rate | 0.0183 | Derived by next generation matrix method to yield $R_e$=1.2 without prior immunity (9) |
| $\gamma^A$ : recovery rate on asymptomatic compartment[a] | 0.1587 | (10) |
| $\gamma^Y$ : recovery rate on symptomatic non-treated compartment | 0.25 | (10) |
| $\tau$ : symptomatic proportion (%) | 57 | (11) |
| $\sigma$ : exposed rate | 0.3448 | (12) |
| $P$: proportion of infections occurring in pre-symptomatic period (%) | 44 | (10) |
| $\omega^P$ : relative infectiousness of infectious individuals in compartment P | 1.3669 | $\omega^P = (\frac{YHR}{\eta} + \frac{1-YHR}{\gamma^Y})\frac{\omega^Y \rho P}{1-P}$ |
| $\omega^A$ : relative infectiousness of infectious individuals in compartment $I^A$ | 0.67 | (10) |
| $\rho$ : symptom onset rate | 0.43478 | 2.3 day average pre-symptomatic period (10) |
| *IFR*: age-stratified infection fatality ratio (%) | Overall: [0.0016, 0.00495, 0.08428, 1.00011, 3.37149]<br>Low risk: [0.00137, 0.00386, 0.06334, 0.60254, 1.73687]<br>High risk: [0.00412, 0.01157, 0.19001, 1.80762, 5.2106] | Age adjusted from Verity et al. (13) |
| *YFR*: age-stratified symptomatic fatality ratio (%) | Overall: [0.00281, 0.00868, 0.14785, 1.75458, 5.9149]<br>Low risk: [0.00241, 0.00677, 0.11112, 1.05709, 3.04713]<br>High risk: [0.00722, 0.0203 , 0.33336, 3.17127, 9.1414] | $YFR = \frac{IFR}{\tau}$ |
| $h$ : high-risk proportion, age specific (%) | [8.2825, 14.1121, 16.5298, 32.9912, 47.0568] | Based on CDC's list of high risk conditions for severe influenza; see Section 4 below (14–31) |
| $rr$ : relative risk of hospitalization for high risk versus low risk individuals in a given age group | 3 | Suggested by CDC for COVID-19 scenario projections (32) and consistent with COVID-19 risk factor studies (33) |

[a]We assume that the duration of the infectious period is the same for asymptomatic and symptomatic cases.

**Table A2.3. Hospitalization parameters**

| **Parameters** | **Value** | **Source** |
|---|---|---|
| $\gamma^H$ : recovery rate in hospitalized compartment | 0.0935 | Austin admissions and discharge data (Avg=10.96. 95% CI = 9.37 to 12.76) (34,35) |
| *YHR*: symptomatic case hospitalization rate (%) | Overall: [ 0.07018, 0.07018, 4.73526, 16.32983, 25.54183]<br>Low risk: [0.0602 , 0.05473, 3.55875, 9.83829, 13.15819]<br>High risk: [ 0.18061, 0.16419, 10.67625, 29.51487, 39.47457] | Age adjusted from Verity et al. (13) |
| $\pi$ : rate of symptomatic individuals go to hospital, age-specific | Low risk: [0.0009, 0.0008, 0.0516, 0.1386, 0.1827]<br>High risk: [0.0027, 0.0024, 0.1499, 0.3818, 0.4903] | $\pi = \frac{\gamma^Y \cdot YHR}{\eta+(\gamma^Y-\eta)YHR}$ |
| $\eta$ : rate from symptom onset to hospitalized | 0.1695 | 5.9 day average from symptom onset to hospital admission Tindale et al. (36) |
| $\mu$ : rate from hospitalized to death | 0.1235 | Austin admissions and discharge data (Avg=7.8, 95% CI = 5.21 to 10.09) (34,35) |
| *HFR*: hospitalized fatality ratio, age specific (%) | [4, 12.365, 3.122, 10.745, 23.158] | $HFR = \frac{IFR}{YHR\tau}$ |
| $\nu$ : death rate on hospitalized individuals, age specific | 0.0617 | $\nu = \frac{\gamma^H HFR}{\mu+(\gamma^H-\mu)HFR}$ |

**Table A2.4. Vaccine parameters**

| Parameters | Value | Source |
|---|---|---|
| $ve^f$: efficacy of full course vaccination, infection-blocking or symptom blocking | After 1st and before 2nd dose:<br>$ve_1^f \sim Triangular(0.295, 0.524, 0.684)$<br>After 2nd dose:<br>$ve_2^f \sim Triangular(0.898, 0.948, 0.976)$ | (37) |
| $ve^s$: vaccine efficacy - single dose only, infection-blocking or symptom blocking | After single dose only:<br>$\sim Triangular(0.756, 0.82, 0.869)$ | (37) |
| $ve^{h,f}$: efficacy of full course vaccination, hybrid model | Efficacy against infection<br>• after 1st and before 2nd dose, $ve_{1,i}^{h,f}$ =0.524<br>• after 2nd dose, $ve_{2,i}^{h,f} = 0.67$<br>Efficacy against symptomatic diseases<br>• after 1st and before 2nd dose, $ve_{1,y}^{h,f}$ =0<br>• after 2nd dose,<br>$ve_{2,y}^{h,f} \sim Triangular(0.691, 0.82, 0.927)$ | (37,38) |
| $ve^{h,s}$: vaccine efficacy - single dose only, hybrid model | Efficacy against infection: $ve_i^{h,s}$ =0.67<br>Efficacy against symptomatic infection:<br>$ve_y^{h,s} \sim Triangular(0.261, 0.455, 0.603)$ | (37,38) |
| *vu*: vaccine uptake rate | 0.5, 0.7, 0.9 | 0.5 is based on a survey conducted by PEW Research Center in September (2); 0.7 is based on a survey conducted by PEW Research Center in May (2); 0.9 is hypothetical |
| $\alpha^{ii}$ : second dose return rate | 0.8 | Assumption |
| Vaccine rollout schedule | 10M nationwide per week and adjusted by Austin MSA population | (39) and discussion with CDC |
| $\delta$ : delay between 1st and 2nd dose | $1/\delta$ =28 days | (40) |
| $\delta^i$, $\delta^{ii}$ : rate of acquiring immunity after the first and second dose | $1/\delta^i = 1/\delta^{ii} = 14$ days | (40) |

**Table A2.5. Home contact matrix (daily number contacts by age group at home)**

| | 0-4y | 5-17y | 18-49y | 50-64y | 65y+ |
|---|---|---|---|---|---|
| **0-4y** | 0.5 | 0.9 | 2.0 | 0.1 | 0.0 |
| **5-17y** | 0.2 | 1.7 | 1.9 | 0.2 | 0.0 |
| **18-49y** | 0.2 | 0.9 | 1.7 | 0.2 | 0.0 |
| **50-64y** | 0.2 | 0.7 | 1.2 | 1.0 | 0.1 |
| **65y+** | 0.1 | 0.7 | 1.0 | 0.3 | 0.6 |

**Table A2.6. School contact matrix (daily number contacts by age group at school)**

| | 0-4y | 5-17y | 18-49y | 50-64y | 65y+ |
|---|---|---|---|---|---|
| **0-4y** | 1.0 | 0.5 | 0.4 | 0.1 | 0.0 |
| **5-17y** | 0.2 | 3.7 | 0.9 | 0.1 | 0.0 |
| **18-49y** | 0.0 | 0.7 | 0.8 | 0.0 | 0.0 |
| **50-64y** | 0.1 | 0.8 | 0.5 | 0.1 | 0.0 |
| **65y+** | 0.0 | 0.0 | 0.1 | 0.0 | 0.0 |

**Table A2.7. Work contact matrix (daily number contacts by age group at work)**

| | 0-4y | 5-17y | 18-49y | 50-64y | 65y+ |
|---|---|---|---|---|---|
| **0-4y** | 0.0 | 0.0 | 0.0 | 0.0 | 0.0 |
| **5-17y** | 0.0 | 0.1 | 0.4 | 0.0 | 0.0 |
| **18-49y** | 0.0 | 0.2 | 4.5 | 0.8 | 0.0 |
| **50-64y** | 0.0 | 0.1 | 2.8 | 0.9 | 0.0 |
| **65y+** | 0.0 | 0.0 | 0.1 | 0.0 | 0.0 |

**Table A2.8. Others contact matrix (daily number contacts by age group at other locations)**

| | 0-4y | 5-17y | 18-49y | 50-64y | 65y+ |
|---|---|---|---|---|---|
| **0-4y** | 0.7 | 0.7 | 1.8 | 0.6 | 0.3 |
| **5-17y** | 0.2 | 2.6 | 2.1 | 0.4 | 0.2 |
| **18-49y** | 0.1 | 0.7 | 3.3 | 0.6 | 0.2 |
| **50-64y** | 0.1 | 0.3 | 2.2 | 1.1 | 0.4 |
| **65y+** | 0.0 | 0.2 | 1.3 | 0.8 | 0.6 |

#### Initial Conditions

Initial conditions were derived using a COVID-19 healthcare forecasting model that we developed in a partnership with the city of Austin and use to provide daily transmission and healthcare projections on a public dashboard (5). The forecasting model is almost identical to the model in this study, but without vaccines. Specifically, it is an expanded stochastic SEIR model with eight disease progression compartments, including symptomatic, pre-symptomatic, asymptomatic patients, and hospitalization. The population is divided into five age groups, with different rates of contacts within and between age groups, a high risk category with each age group, and age- and risk-specific rates of hospitalization. The demographic, health and mixing parameters are identical to those assumed in this study.

To make daily dashboard projections, we incorporate anonymized local mobility data from SafeGraph (41) into transmission rate. We assume published estimates for all disease progression parameters and calibrate the remaining unknown states and parameters to local COVID-19 hospital admissions and discharge data using iterated filtering made available through the POMP R package (42). The result of the statistical inference is posterior densities for parameters governing the impact of mobility on transmission and the reporting process of hospitalization data, and for hidden states of the model including the number of infected individuals. Additional details are provided in ref. (43).

To obtain initial conditions for this study, we used the states in the fitted model based on data through November 7, 2020, as given in Table A2.9.

**Table A2.9. Initial states of model compartments.** Values indicate the number of individuals in each age-risk group compartment at the start of the simulations.

| Age | Risk | S | E | $I^A$ | $I^P$ | $I^Y$ | $I^H$ | R | D |
|---|---|---|---|---|---|---|---|---|---|
| **0-4 yr** | **High** | 8809 | 7 | 5 | 3 | 5 | 0 | 524 | 0 |
| | **Low** | 121035 | 95 | 87 | 42 | 73 | 0 | 7190 | 0 |
| **5-17 yr** | **High** | 33884 | 40 | 39 | 21 | 33 | 0 | 3418 | 0 |
| | **Low** | 296425 | 372 | 343 | 169 | 279 | 0 | 29580 | 1 |
| **18-49 yr** | **High** | 142443 | 165 | 143 | 70 | 140 | 35 | 13158 | 26 |
| | **Low** | 834952 | 949 | 884 | 442 | 746 | 21 | 77836 | 16 |
| **50-64 yr** | **High** | 100333 | 102 | 84 | 44 | 87 | 42 | 7262 | 255 |
| | **Low** | 231158 | 219 | 201 | 97 | 171 | 9 | 17370 | 57 |
| **65+ yr** | **High** | 100101 | 45 | 40 | 21 | 43 | 20 | 3261 | 236 |
| | **Low** | 127788 | 60 | 55 | 26 | 51 | 2 | 4485 | 31 |

### Section 3. Sensitivity Analyses

**Sensitivity analysis with respect to the effective reproduction number (*$R_e$*) of the virus in November 8, 2021**

Our base scenarios assume an effective reproductive number ($R_e$) of 1.2 (5). Here, we provide projections based on three alternative transmission rate scenarios: $R_e$= 1.5, $R_e$= 1.05, and $R_e$ increasing linearly from 1.2 to 1.8 by May 1, 2021 and remaining at 1.8 through September 17, 2021.

Under the lowest transmission rate scenario ($R_e$= 1.05), the pace of the epidemic is slower and the expected impact of vaccinations on overall mortality and the duration of the pandemic is greater than in the base case (Figure A3.1). Under the moderate transmission scenario ($R_e$= 1.5), the reverse occurs. The pandemic sweeps through and achieves herd immunity rapidly, leaving little opportunity for vaccines to prevent infections and deaths.

The increasing transmission rate scenario roughly models the emergence and rapid spread of a more transmissible SARS-CoV-2 variant, like B.1.1.7. The pandemic wave reaches a peak at a similar time to the base case, but reaches a much higher prevalence of COVID-19. Overall, vaccination has a lower impact on COVID-19 mortality and the duration of the pandemic wave, but risk-based prioritization and community uptake have similar relative impacts.

Consider the best of the vaccination scenarios—90% uptake of a perfectly-prioritized rollout of a vaccine that reduces susceptibility beginning on January 15, 2021. The expected COVID-19 deaths averted between January 15 and September 17, 2021 are 60% (95% CI: 51%-67%), 56% (95% CI: 51%-60%), 22% (95% CI: 17%-25%), and 41% for the $R_e$= 1.05, $R_e$= 1.2, $R_e$= 1.5, and dynamic $R_e$ scenarios, respectively. The expected numbers of COVID-19 deaths per 100,000 during the peak week are 27 (95% CI: 22-33), 107 (95% CI: 100-122), 386 (95% CI: 370-407), and 439 (95% CI: 413-474), respectively.

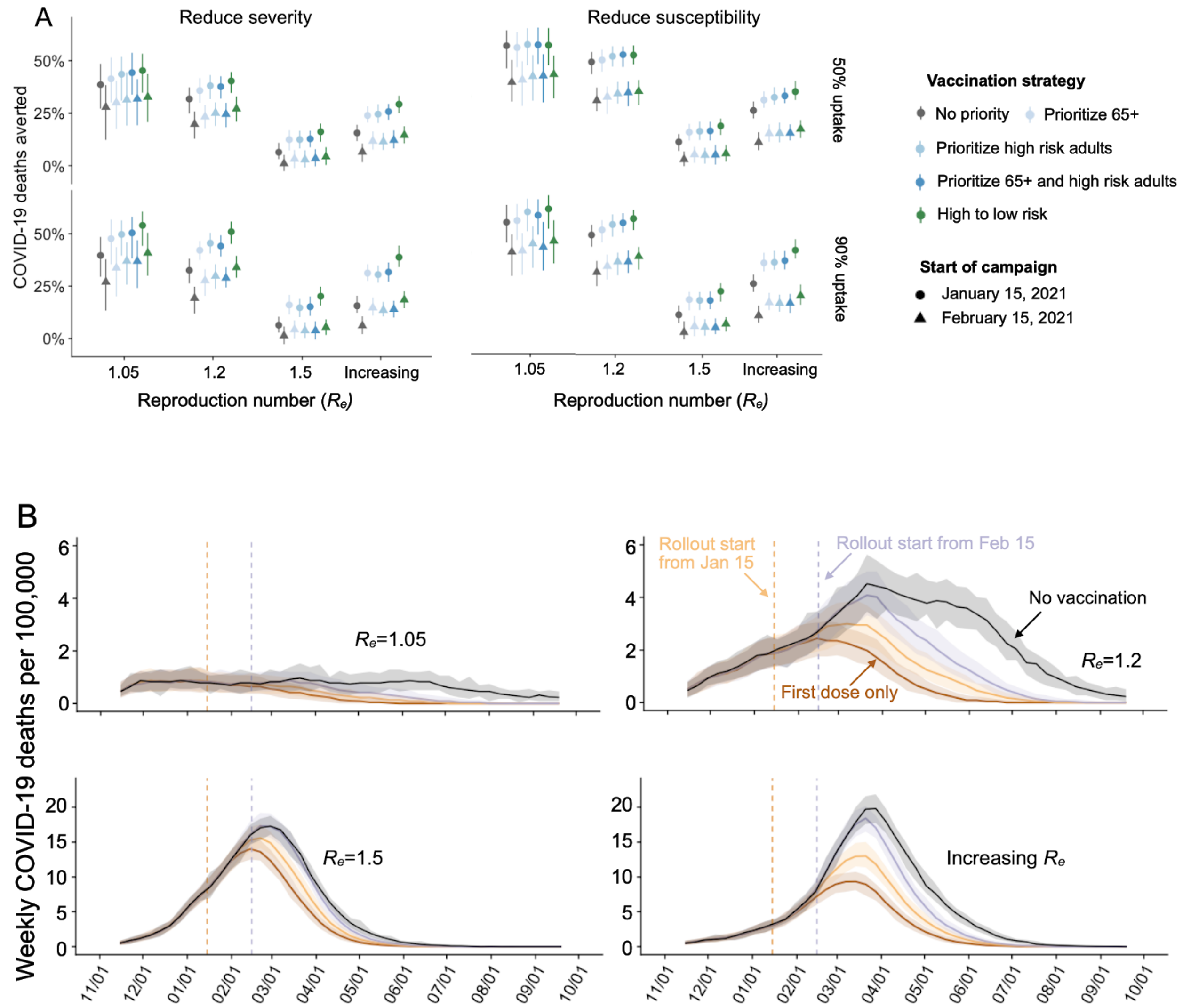


**Figure A3.1. Projected COVID-19 mortality in the Austin-Round Rock MSA from November 8, 2020 to September 17, 2021 under different vaccination scenarios, assuming either $R_e$ = 1.05, $R_e$ = 1.2, $R_e$ = 1.5, or $R_e$ increases linearly from 1.2 to 1.8 by May 1, 2021 and remains at 1.8 thereafter.** A) COVID-19 deaths averted after January 15, 2021 under combinations of: vaccine uptake, either 50% (top) or 90% (bottom); type of protection, either symptom blocking (left) or infection blocking (right); rollout dates, either January 15 (circles) or February 15 (triangles); and risk prioritization, either no priority (gray), prioritize all adults over 65y (light blue), adults with high-risk comorbidities (medium blue), or the combination of the two (dark blue), or a ten-phase risk-ordered strategy (green) that sequentially vaccinates >65y high risk, 50-64y high risk, >65y low risk, 18-49y high risk, 50-64y low risk, 18-49y low risk, 0-4y high risk, 5-17y high risk, 0-4y low risk, 5-17y low risk. Points and whiskers indicate the median and 95% CI across 200 paired stochastic simulations. B) Weekly incident COVID-19 deaths per 100,000 assuming intermediate (70%) uptake (2) without vaccine (black) or under a ten-phase risk-based rollout of a 95% efficacious infection-blocking, starting either January 15 (orange) or February 15 (purple), for the indicated $R_e$ scenario. The brown lines assume that only first doses are administered starting January 15. Solid lines and shading indicate the median and 95% CI across 200 stochastic simulations, respectively.

**Sensitivity analysis with respect to prior immunity and the prevalence of infections as of November 8, 2020**

Our base scenarios assume that 7.6% of the individuals in the population have obtained immunity due to prior infection by early November and that 0.3% of the population is infected on the first day of the simulation (November 8, 2020) (Table A2.9) (5). In Figure A3.2, we provide projections assuming that twice as many people are infected (0.6%) are infected at the start of each simulation or twice as many people (15.2%) are immune at the outset. All projections assume the same transmission rate per contact, which was derived to produce a reproduction number of $R_e$=1.2 in the absence of prior immunity (Table A2.2).

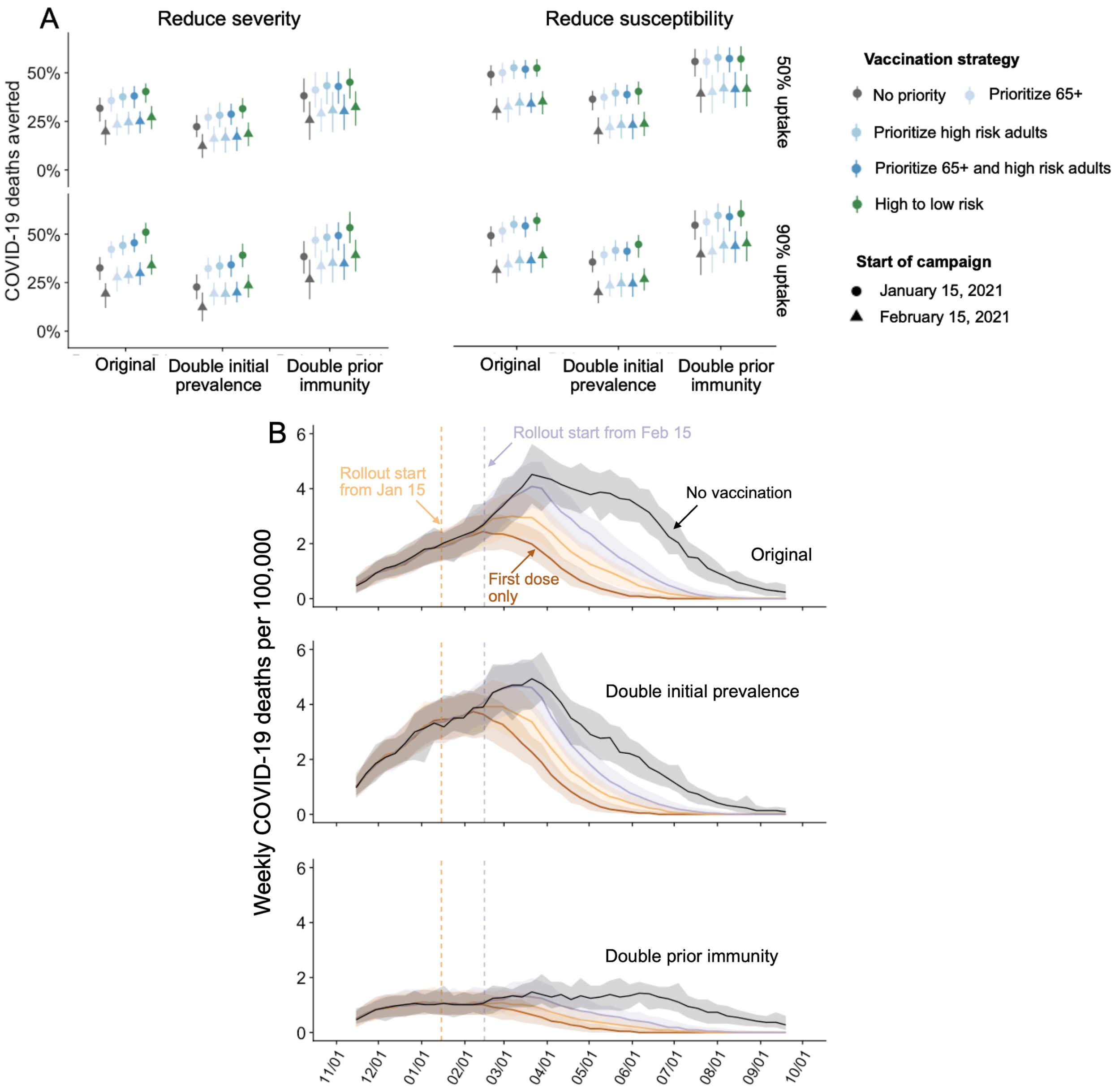


**Figure A3.2. Projected COVID-19 mortality in the Austin-Round Rock MSA from November 8, 2020 to September 17, 2021 under different vaccine rollout scenarios, assuming a higher initial prevalence (0.6%) or prior immunity (15.2%).** A) COVID-19 deaths averted after January 15, 2021 under combinations of: vaccine uptake, either 50% (top) or 90% (bottom); type of protection, either symptom blocking (left) or infection blocking (right); initial conditions (x-axis); rollout dates, either January 15 (circles) or February 15 (triangles); and risk prioritization (colors). Points and whiskers indicate the median and 95% CI across 200 paired stochastic simulations. B) Weekly incident COVID-19 deaths per 100,000 assuming intermediate (70%) uptake (2) without vaccine (black) or under a ten-phase risk-based rollout of a 95% efficacious infection-blocking, starting either January 15 (orange) or February 15 (purple), under the original scenario (top), double initial prevalence (middle), and double prior immunity (bottom). The brown line assumes that only first doses are administered starting January 15. Solid lines and shading indicate the median and 95% CI across 200 stochastic simulations, respectively.

**Sensitivity analysis with respect to vaccine efficacy**

Our original analyses assume 95% vaccine efficacy following two doses and 80% efficacy following a single dose (Table A2.4). However, recent studies have raised concerns that some SARS-CoV-2 vaccines may have reduced efficacy against emerging and future variants, including B.1.351 (44). In Figure A3.3, we provide projections assuming that the vaccine efficacy is reduced by 50%.

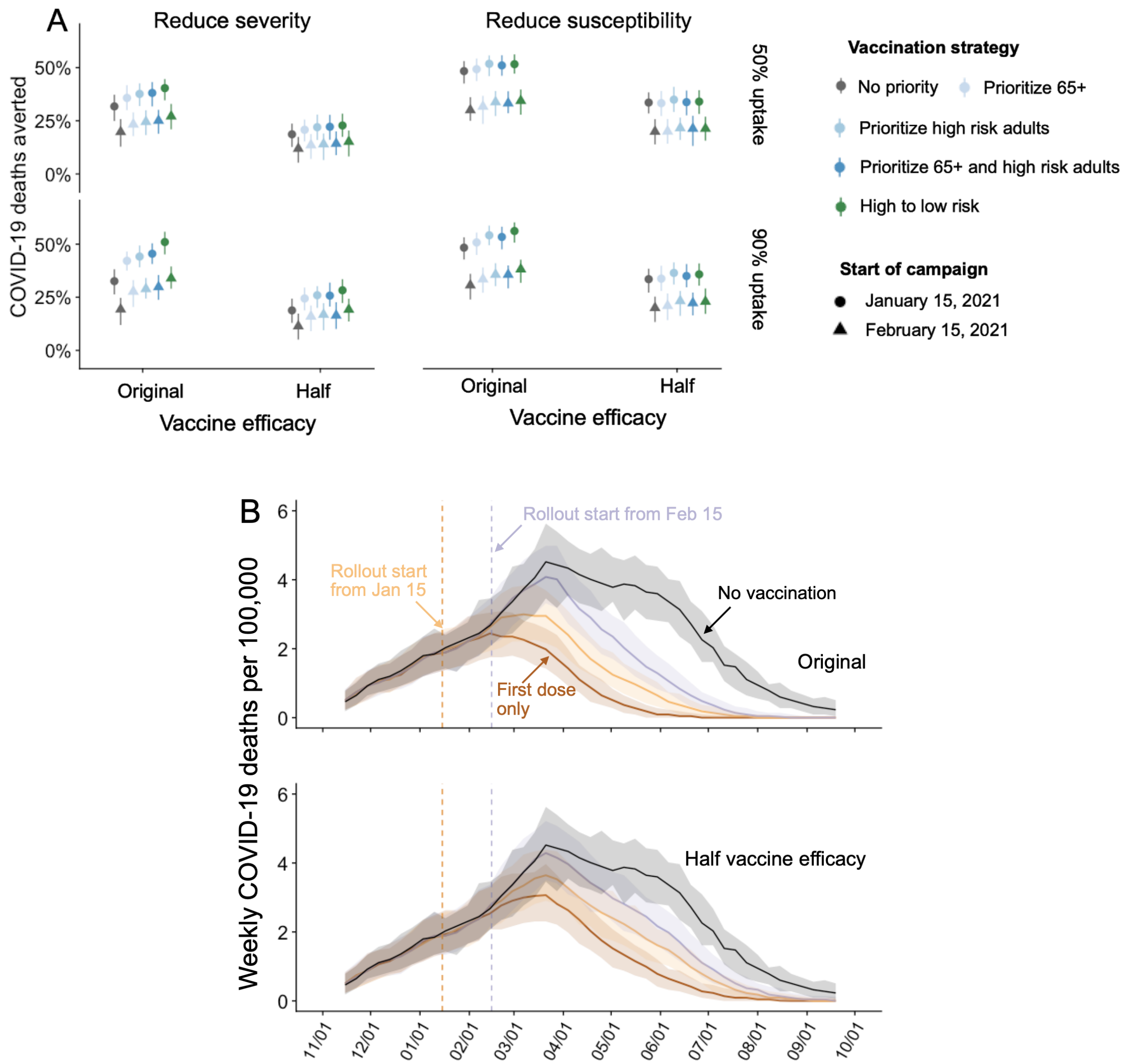


**Figure A3.3. Projected COVID-19 mortality in the Austin-Round Rock MSA from November 8, 2020 to September 17, 2021 under different vaccine rollout scenarios, assuming a 50% reduction in vaccine efficacy.** A) COVID-19 deaths averted after January 15, 2021 under combinations of: vaccine uptake, either 50% (top) or 90% (bottom); type of protection, either symptom blocking (left) or infection blocking (right); vaccine efficacy, either original or half of original (x-axis); rollout dates, either January 15 (circles) or February 15 (triangles); and risk prioritization (colors). Points and whiskers indicate the median and 95% CI across 200 paired stochastic simulations. B) Weekly incident COVID-19 deaths per 100,000 assuming intermediate (70%) uptake (2) without vaccine (black) or under a ten-phase risk-based rollout of a 95% efficacious infection-blocking, starting either January 15 (orange) or February 15 (purple) with original vaccine efficacy (top) or 50% lower vaccine efficacy (bottom). The brown line assumes that only first doses are administered starting January 15. Solid lines and shading indicate the median and 95% CI across 200 stochastic simulations, respectively.

### Section 4. Estimation of age-stratified proportion of population at high-risk for COVID-10 complications

We estimate age-specific proportions of the population at high risk of complications from COVID-19 based on data for Austin, TX and Round-Rock, TX from the CDC's 500 cities project (Figure A4) (17). We assume that high risk conditions for COVID-19 are the same as those specified for influenza by the CDC (14). The CDC's 500 cities project provides city-specific estimates of prevalence for several of these conditions among adults (18). The estimates were obtained from the 2015-2016 Behavioral Risk Factor Surveillance System (BRFSS) data using a small-area estimation methodology called multi-level regression and poststratification (16,20). It links geocoded health surveys to high spatial resolution population demographic and socioeconomic data (16).

##### Estimating high-risk proportions for adults

To estimate the proportion of adults at high risk for complications, we use the CDC's 500 cities data, as well as data on the prevalence of HIV/AIDS, obesity and pregnancy among adults (Table A7).

The CDC 500 cities dataset includes the prevalence of each condition on its own, rather than the prevalence of multiple conditions (e.g., dyads or triads). Thus, we use separate co-morbidity estimates to determine overlap. Reference about chronic conditions (19) gives US estimates for the proportion of the adult population with 0, 1 or 2+ chronic conditions, per age group. Using this and the 500 cities data we can estimate the proportion of the population pHR in each age group in each city with at least one chronic condition listed in the CDC 500 cities data (Table A7) putting them at high-risk for flu complications.

HIV: We use the data from table 20a in CDC HIV surveillance report (21) to estimate the population in each risk group living with HIV in the US (last column, 2015 data). Assuming independence between HIV and other chronic conditions, we increase the proportion of the population at high-risk for influenza to account for individuals with HIV but no other underlying conditions.

Morbid obesity: A BMI over 40kg/m$^2$ indicates morbid obesity, and is considered high risk for influenza. The 500 Cities Project reports the prevalence of obese people in each city with

BMI over 30kg/m$^2$ (not necessarily morbid obesity). We use the data from table 1 in Sturm and Hattori (22) to estimate the proportion of people with BMI>30 that actually have BMI>40 (across the US); we then apply this to the 500 Cities obesity data to estimate the proportion of people who are morbidly obese in each city. Table 1 of Morgan et al. (23) suggests that 51.2% of morbidly obese adults have at least one other high risk chronic condition, and update our high-risk population estimates accordingly to account for overlap.

Pregnancy: We separately estimate the number of pregnant women in each age group and each city, following the methodology in CDC reproductive health report (24). We assume independence between any of the high-risk factors and pregnancy, and further assume that half the population are women.

**Estimating high-risk proportions for children**

Since the 500 Cities Project only reports data for adults 18 years and older, we take a different approach to estimating the proportion of children at high risk for severe influenza. The two most prevalent risk factors for children are asthma and obesity; we also account for childhood diabetes, HIV and cancer.

From Miller et al. (25), we obtain national estimates of chronic conditions in children. For asthma, we assume that variation among cities will be similar for children and adults. Thus, we use the relative prevalences of asthma in adults to scale our estimates for children in each city. The prevalence of HIV and cancer in children are taken from CDC HIV surveillance report (21) and cancer research report (26), respectively.

We first estimate the proportion of children having either asthma, diabetes, cancer or HIV (assuming no overlap in these conditions). We estimate city-level morbid obesity in children using the estimated morbid obesity in adults multiplied by a national constant ratio for each age group estimated from Hales et al. (27), this ratio represents the prevalence in morbid obesity in children given the one observed in adults. From Morgan et al. (23), we estimate that 25% of morbidly obese children have another high-risk condition and adjust our final estimates accordingly.

**Resulting estimates**

We compare our estimates for the Austin-Round Rock Metropolitan Area to published national-level estimates (28) of the proportion of each age group with underlying high risk conditions (Table A8). The biggest difference is observed in older adults, with Austin having a lower proportion at risk for complications for COVID-19 than the national average; for 25-39 year olds the high risk proportion is slightly higher than the national average.

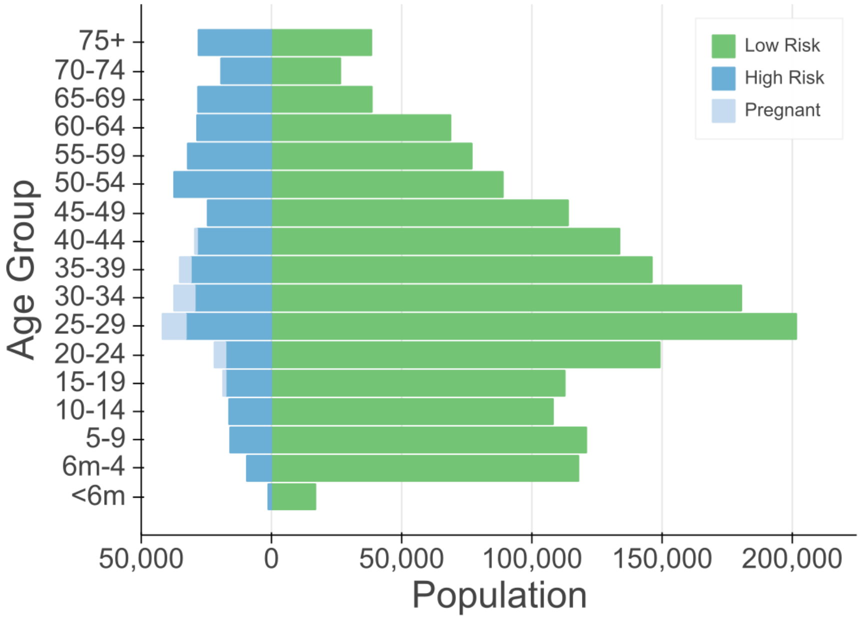


**Figure A4. Demographic and risk composition of the Austin-Round Rock MSA.** Bars indicate age-specific population sizes, separated by low risk, high risk, and pregnant. High risk is defined as individuals with cancer, chronic kidney disease, COPD, heart disease, stroke, asthma, diabetes, HIV/AIDS, and morbid obesity, as estimated from the CDC 500 Cities Project (17), reported HIV prevalence (21) and reported morbid obesity prevalence (22,23), corrected for multiple conditions. The population of pregnant women is derived using the CDC's method combining fertility, abortion and fetal loss rates (29–31).

**Table A4.1 High-risk conditions for influenza and data sources for prevalence estimation**

| Condition | Data source |
|---|---|
| Cancer (except skin), chronic kidney disease, COPD, coronary heart disease, stroke, asthma, diabetes | CDC 500 cities (17) |
| HIV/AIDS | CDC HIV Surveillance report (18) |
| Obesity | CDC 500 cities (17), Sturm and Hattori (22), Morgan et al. (23) |
| Pregnancy | National Vital Statistics Reports (29) and abortion data (30) |

**Table A4.2. Comparison between published national estimates and Austin-Round Rock MSA estimates of the percent of the population at high-risk of influenza/COVID-19 complications.**

| **Age Group** | **National estimates** (27) | **Austin (excluding pregnancy)** | **Pregnant women (proportion of age group)** |
|---|---|---|---|
| 0 to 6 months | NA | 6.8 | - |
| 6 months to 4 years | 6.8 | 7.4 | - |
| 5 to 9 years | 11.7 | 11.6 | - |
| 10 to 14 years | 11.7 | 13.0 | - |
| 15 to 19 years | 11.8 | 13.3 | 1.7 |
| 20 to 24 years | 12.4 | 10.3 | 5.1 |
| 25 to 34 years | 15.7 | 13.5 | 7.8 |
| 35 to 39 years | 15.7 | 17.0 | 5.1 |
| 40 to 44 years | 15.7 | 17.4 | 1.2 |
| 45 to 49 years | 15.7 | 17.7 | - |
| 50 to 54 years | 30.6 | 29.6 | - |
| 55 to 60 years | 30.6 | 29.5 | - |
| 60 to 64 years | 30.6 | 29.3 | - |
| 65 to 69 years | 47.0 | 42.2 | - |
| 70 to 74 years | 47.0 | 42.2 | - |
| 75 years and older | 47.0 | 42.2 | - |

https://ir.novavax.com/news-releases/news-release-details/novavax-covid-19-vaccine-demonstrates-8
93-efficacy-uk-phase-3
